# Characteristics and outcomes of cases of children and adolescents with pediatric inflammatory multisystem syndrome in a tertiary care center in Mexico City

**DOI:** 10.1101/2021.12.23.21268188

**Authors:** Ricardo Gil Guevara, María de Lourdes Marroquín Yáñez, Rodolfo Norberto Jiménez-Juárez, Víctor Olivar Lopez, Adrián Chávez Lopez, Juan José Luis Sienra Monge, Lourdes Maria del Carmen Jamaica Balderas, Silvia Alexandra Martínez Herrera, Clemen Domínguez-Barrera, Julio Erdmenger Orellana, Horacio Márquez González, Miguel Klünder-Klünder, Jaime Nieto Zermeño, Mónica Villa Guillen, Nadia González García, Maria F Castilla-Peon

**Author notes:** These authors have contributed equally to this work and share first authorship (Ricardo Gil Guevara, María de Lourdes Marroquín Yáñez). These authors have contributed equally to this work and are both corresponding authors (Nadia González García, Maria F Castilla-Peon). **Correspondence**: Maria F. Castilla-Peon, Nadia Gonzalez-García.

## Abstract

**Background:** pediatric inflammatory multisystem syndrome (PIMS) is a complication of severe acute respiratory syndrome coronavirus 2 (SARS-CoV-2) infection in children that resembles Kawasaki syndrome and places them at high risk of cardiorespiratory instability and/or cardiac damage. This study aims to describe the clinical presentation and outcomes of patients with PIMS in Mexico City.

**Methods:** This was an observational study of children hospitalized for PIMS based on the Centers for Disease Control and Prevention case definition criteria, in a single tertiary care pediatric center in Mexico City between May 1, 2020, and September 30, 2021. Demographic characteristics, epidemiological data, medical history, laboratory tests, cardiology evaluations, treatment, and clinical outcomes were analyzed.

**Results:** Seventy-five cases fulfilled the case definition criteria for PIMS (median age: 10.9 years, Interquartile range [IQR]: 5.6–15.6). Fifteen (20%) patients had a severe underlying disease, 48 (64%) were admitted to the intensive care unit, 33 (44%) required invasive mechanical ventilation and 39 (52%) received vasopressor support. The patients were clustered through latent class analysis based on identified symptoms: Cluster 1 had rash or gastrointestinal symptoms (n = 60) and cluster 2 were those with predominantly respiratory manifestations (n = 15). Two patients (2.7%) died, and both had severe underlying conditions. Five patients (6.7%), all from cluster 1, developed coronary aneurysms.

**Conclusion:** There were a high proportion of patients with severe respiratory involvement and positive RT-PCR SARS-CoV-2 and very few cases of coronary aneurysms in our study which suggests that a high proportion of the children had severe acute COVID-19. The clinical manifestations and outcomes are comparable to previously reported international studies.

## 1 Introduction

Severe acute respiratory syndrome coronavirus 2 (SARS-CoV-2) infection usually has a mild clinical presentation in children. However, in rare cases, children can be severely affected and have clinical manifestations, different from adults. In April 2020, some reports described a clinical syndrome similar to Kawasaki disease or toxic shock syndrome temporally associated with current or recent SARS-CoV-2 infection in the pediatric population (1). Since then, this syndrome has been recognized worldwide and named pediatric inflammatory multisystem syndrome (PIMS) or multisystem inflammatory syndrome in children (MIS-C) (2–6).

PIMS case definition varies slightly between different health agencies (7–9) and it is likely to change over time. The criteria include fever, inflammatory marker elevation, multisystem organ involvement, evidence of current or recent SARS-CoV-2 infection, and exclusion of alternative diagnoses. Besides the need for hospitalization for life support, an outcome of concern is myocardial and coronary artery involvement similar to those observed in Kawasaki disease (10–12).

PIMS incidence has been estimated to be approximately 3–5 per 10,000 individuals younger than 21 years of age infected with SARS-CoV-2, and Black and Hispanic ethnicities have been associated with a higher incidence (13). Epidemiological description of PIMS has been difficult because of varying awareness of the condition in different clinical scenarios and because of the diversity of clinical manifestations. Besides, there is an overlap between the clinical manifestations of severe acute COVID-19 and PIMS (10). Hence, the current PIMS case definition is purposely broad and unspecific to obtain maximum and comprehensive information about this phenomenon. However, increasing evidence suggests the existence of different phenotypes of PIMS, the more clearly defined of them being a Kawasaki-like syndrome, a toxic shock-like syndrome, and a predominantly respiratory syndrome which might have different pathogenesis and clinical outcomes (14–16).

This study aims to describe the clinical characteristics and outcomes of PIMS cases admitted to a tertiary care pediatric center in Mexico City. **A secondary aim was to define potential clinical subgroups of children with PIMS**.

## 2 Materials and Methods

### 2.1 Study design

This was **an observational prospective** study of PIMS cases diagnosed in ‘Hospital Infantil de Mexico Federico Gómez’ (Federico Gómez Mexico Children’s Hospital), a tertiary care pediatric facility officially designated to treat patients with severe SARS-CoV-2 infection who are less than 18 years old, and without public or private health insurance in Mexico City.

Probable PIMS cases were identified in the emergency room, transferred from other health care facilities directly to the intensive care unit, or identified by treating physicians in the general hospitalization ward for COVID-19 patients. Cases were evaluated to determine if they met the Centers for Disease Control and Prevention (CDC) PIMS case definition (7). This case definition included the presence of fever, the elevation of inflammatory markers, signs of involvement of at least two organ systems requiring hospitalization, and evidence of recent SARS-CoV-2 exposure or infection. Patients were excluded if they had another plausible explanation for the illness. When there was uncertainty about an alternative diagnosis, the complete case file was reviewed by an expert panel which included an infectious disease specialist and a critical care specialist, until a consensus was reached. Detailed inclusion criteria are specified in Chart 1.

### 2.2 Data collection

Clinical and laboratory data were extracted from the patients’ clinical file and included demographics, underlying medical conditions, clinical manifestations, laboratory values, management, and outcomes. For laboratory values with more than one measurement, the worst value measured within the first three days of hospitalization was obtained. Vital support management (respiratory support and vasopressor utilization), intravenous immune globulin, systemic steroids, and anticoagulant therapy were documented as the main therapeutic interventions. Outcomes of interest were intensive care unit (ICU) admission, length of ICU stay and hospitalization, need for invasive mechanical ventilation, vasopressor support, myocardial depression (left ventricular ejection fraction < 55%), coronary aneurysms (coronary artery diameter z-score ≥ 2.5), or dilatation (z-score >2–<2.5) on the echocardiographic evaluation performed during hospitalization, and in-hospital mortality. Markers with more than 20% missing data were not analyzed.

For this study, severe respiratory involvement was defined as infiltrates on chest X-ray or computed tomography plus the need for oxygen administration with a non-rebreathable mask or a higher oxygen concentration device. The variable ‘gastrointestinal symptoms’ was defined as the presence of at least one of the following: abdominal pain, diarrhea, or emesis.

The study center has an important regular population of patients with severe underlying diseases. Hence, to have a picture more alike general population, we performed a sub-analysis with the exclusion of cases with severe underlying diseases (i.e., cancer and other forms of immunosuppression diseases, neuromuscular disability, chronic respiratory disease except for asthma, congenital cardiopathies, and chronic kidney failure).

### 2.3 Statistical analysis

Descriptive analysis was conducted using STATA v.14.0 (StataCorp, Tx). Categorical variables were reported as frequencies and continuous variables as medians and interquartile ranges (IQR). Analysis was performed on the whole sample and three subgroups. In the first subgroup, cases with severe underlying conditions were excluded. The other two subgroups were identified using latent class analysis (LCA). Two class LCA were conducted using the R software package ‘poLCA’ (17) with 100 iterations to identify the clusters. The indicator variables used in LCA were the presence or absence of severe respiratory involvement, rash and gastrointestinal symptoms. The fit of each model was assessed using a Bayesian Information Criterion score.

The study was approved by the ethics review board of Federico Gómez Mexico Children’s Hospital (Reg: HIM2020-031), including the publication of de-identified data.

## 3 Results

Reports of one hundred and sixty-seven (167) probable cases of PIMS admitted to the hospital between May 1, 2020, and September 30, 2021, were sent by clinical departments. For reference, during this period, 8,193 real-time polymerase chain reaction (RT-PCR) tests for SARS-CoV-2 were performed in the study center, which yielded 837 positive results. **The temporal distribution of cases follows the pattern of the three epidemic peaks in Mexico City during the study period (Figure 1)**.

**Figure 1.**
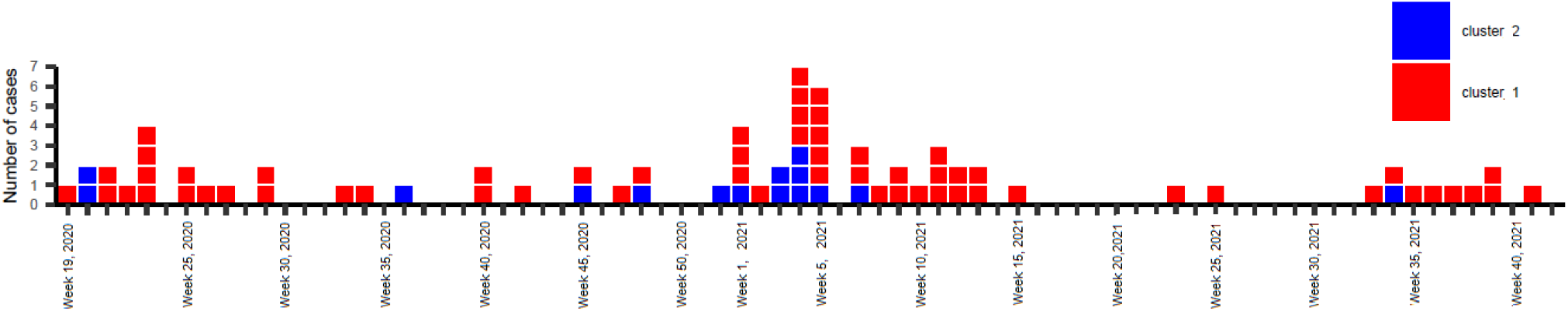
Temporal distribution of PIMS cases by cluster from May 1^st^, 2020 (Week 19, 2020) to September 30, 2021 (Week 40, 2021).

From the 167 received reports, 42 were from the emergency department, 34 from the ICU, and 91 from the infectious disease department. Duplicated reports were eliminated (n = 23) and 23 reports failed to meet inclusion criteria. **Severe conditions were not unusual among patients being attended at the study center. Besides, testing for SARS-CoV-2 is performed routinely before hospital admission. Hence, the scenario of SARS-CoV-2 infection in a severely ill patient was common in the received reports. Forty-six cases were excluded because either elevated inflammation markers or system failures had an obvious cause other than the exposure to SARS-CoV-2. Of these, in 13 cases an infectious process other than SARS-CoV-2 was identified, such as acute complicated appendicitis (n=5), positive blood cultures (n=3), neck abscess (n=2), aspergillosis (n=1), neutropenic colitis (n=1), and influenza coinfection (n=1). In 6 cases, lymphopenia and/or thrombocytopenia were attributed to drugs, mainly antineoplastic chemotherapy. Five cases presented with a new onset malignancy, two had a reactivation of systemic lupus erythematosus, one had a macrophage activation syndrome associated with idiopathic juvenile arthritis and one case was diagnosed with Kawasaki syndrome (no evidence of SARS-CoV-2 exposure was found in this case). Other excluded cases which did not meet criteria for PIMS aside from the organic failure attributed to the comorbid condition, had congenital cardiopathies (n=5), neurologic conditions (n=3), chronic renal disease (n=2), ketoacidosis (n=2), aplastic anemia, digoxin intoxication, post cardiorespiratory arrest status, histiocytosis, intestinal occlusion, and a vascular neoplasm which originated consumption coagulopathy. Of the 46 excluded cases, only the one diagnosed with Kawasaki syndrome received intravenous immune globulin (IVIG)**.

**Among the 75 included cases there was only one case presenting with clinical manifestations of acute abdomen and was operated in the referring health care center. This case was transferred to the study center because of his poor evolution after surgery and we do not have information on the surgical findings. Other cases presenting with acute abdomen (n=5) were not included in the analysis because surgical findings explained the inflammatory clinical manifestations**.

The results for the 75 included cases are as follow: the median (interquartile range [IQR]) age was 10.9 (5.6-15.6) years and more than half (52%) were females. A total of 45.3% of the participants had at least one underlying condition, including obesity (25%), cancer (9%), neuromuscular (8%), and respiratory (7%) diseases (Table 1).

**Table.1.** Clinical presentation of cases with PIMS.

|  | Total population<br>(N=75)<sup>1</sup> | Without serious<br>underlying<br>disease<sup>2</sup><br>(N=60)<sup>1</sup> | Cluster 1<br>With GIS<sup>3</sup> or<br>Rash<br>(N=60)<sup>1</sup> | Cluster 2<br>Predominant respiratory<br>(N=15)<sup>1</sup> | p * |
| --- | --- | --- | --- | --- | --- |
| Female sex -no. (%) | 39 (52%) | 33(55%) | 33 (55%) | 6 (40%) | 0.23 |
| Age - median (IQR) | 10.9 (5.6-15.6) | 10.6 (5.5-15.0) | 10.3 (5.7-13.5) | 15.3 (5.4-16.6) | 0.08 |
| Age group - n (%) |  |  |  |  |  |
| 4<1 y | 5 (6.7%) | 5 (8.3%) | 4(6.7%) | 1 (6.7%) |  |
| 1-4 y | 11 (17.3%) | 9 (15%) | 9 (15%) | 2 (1.3%) |  |
| 5-9 y | 13 (17.1%) | 11 (18.3%) | 12 (0.2%) | 1 (6.7%) | 0.12 |
| 10-14 y | 26 (34.7%) | 21 (35%) | 23 (38.3%) | 3 (20%) |  |
| 15-18 y | 20 (26.7%) | 14 (23.3%) | 12 (20%) | 8 (53%) |  |
| Days of symptoms before admission | 7(4-9) | 7 (5-9) | 6 (4-8) | 7 (1-9) |  |
| Symptom's onset posterior to June 1, 2021 | 11 (14.7%) | 10 (16.7%) | 10 (16.7%) | 1 (6.7%) | 0.30 |
| SARS-COV2 testing, No. (%) |  |  |  |  |  |
| RT- PCR positive | 57/ 71 (80%) | 43/ 56 (76.8%) | 44/58 (75.9%) | 13/13 (100%) | 0.04 |
| Serology positive | 25/ 41 (61%) | 25/46 (54%) | 25/ 36(69%) | 0/5* | 0.01 |
| Underlying medical conditions - no. (%) |  |  |  |  |  |
| Obesity (BMI > 95<sup>o</sup>percentile) | 18 (24%) | 14 (23.0%) | 12 (20%) | 6 (40%) | 0.10 |
| Respiratory | 5 (6.7%) | 3 (5%) | 2 (3.3%) | 3(20%) | 0.05 |
| Neuromuscular | 6 (8.0%) | 0 | 2 (3.3%) | 4 (26.7%) | 0.01 |
| Cardiovascular | 3 (4%) | 0 | 2 (3.3%) | 1 (6.7%) | 0.49 |
| Cancer | 7 (9.3%) | 0 | 2 (3.3%) | 5 (33%) | 0.003 |
| Other, immunosuppression | 2 (3%) | 0 | 1 (1.7%) | 1 (6.7%) | 0.32 |
| Other comorbidity | 10 (13.3%) | 6 (10%)<sup>4</sup> | 8 (13.3%) | 2 (13.3%) | 0.64 |
| At least 1 underlying condition | 34 (45.3%) | 19 (31.2%) | 22 (36.7%) | 12 (80%) | 0.003 |
| Clinical manifestations – no. (%) |  |  |  |  |  |
| Cough | 39(52%) | 32 (53.3%) | 33 (55%) | 6 (40%) | 0.23 |
| Rash | 36 (48%) | 33 (55%) | 36 (60%) | 0 | <0.001 |
| Conjunctivitis | 39 (50.6 %) | 39 (52%) | 36 (60%) | 2 (13.3%) | 0.001 |
| Diarrhea | 33 (44%) | 27 (45%) | 33 (55%) | 0 | <0.001 |
| Emesis | 36 (48%) | 33 (55%) | 36(60%) | 0 | <0.001 |
| Abdominal pain | 38 (51%) | 36 (60%) | 38 (63.3%) | 0 | <0.001 |
| Headache | 34(45.3%) | 25 (41.7%) | 27 (45%) | 7 (46.7%) | 0.57 |
| Rhinorrhea | 24 (32%) | 20 (33.3%) | 19 (31.7%) | 5 (33.2%) | 0.56 |
| Respiratory Distress | 31 (41%) | 24 (40%) | 21 (35%) | 10 (66.7%) | 0.03 |
| Lower respiratory symptoms | 41 (54%) | 31 (51.7%) | 29 (48.3%) | 12 (80%) | 0.03 |
| Anosmia/Dysgeusia | 7 (9.3%) | 6 (10%) | 4 (6.6%) | 3 (20%) | 0.14 |
| Arthralgias/Myalgias | 14(18.7%) | 11 (18.3%) | 13 (21.7%) | 1 (6.7%) | 0.17 |
| Oxygen Saturation<90% | 30 (40%) | 20 (33.3%) | 22 (36.7%) | 8 (53.3%) | 0.19 |
| Laboratory values within 72 h of admission – median (IQR) / n (%) |  |  |  |  |  |
| Neutrophil count x10<sup>6</sup> cells/mcl | 6.1 (1.7-9.2) | 7.1(3.1-10.1) | 6-5 (1.9-9.8) | 3.1(0.5-7.1) | 0.04 |
| Neutrophilia (>7500 cells/mm<sup>3</sup>) | 29/74 (39.2 %) | 27/59 (45.8%) | 27/59 (45.8%) | 2 (13.3%) | 0.02 |
| Neutropenia (<1000 cakes/mm<sup>3</sup>) | 10 (13%) | 6 (10%) | 5/59 (8.5%) | 5 (33%) | 0.02 |
| Lymphocyte count x10<sup>6</sup> cells/mm<sup>3</sup> | 1.5 (0.5-2.4) | 1.6 (0.8-2.1) | 1.6 (0.6-2.8) | 0.8 (0.4-1.8) | 0.16 |
| Lymphopenia<sup>5</sup> | 43/73 (58.9 %) | 33/58 (56.9%) | 33/58 (56.9%) | 10 (66.7%) | 0.35 |
| Platelets x 10<sup>3</sup>cells/mm<sup>3</sup> | 162 (94-250) | 166(110-271) | 162 (105-235) | 162 (43-330) | 0.82 |
| Fibrinogen, mg/dl | 510 (404-668) | 558(431-724) | 516 (412-671) | 510 (361-669) | 0.66 |
| Fibrinogen> 400 mg/dl | 52 (69.3%) | 44/56 (78.6%) | 43 (71.7%) | 10 (76.9%) | 0.5 |
| Ferritin, mcg/L | 617 (407-1370) | 559 (380-1110) | 604 (417-1150) | 1370 (325-6071) | 0.25 |
| Ferritin> 300 mg/dl | 53 (70.1%) | 41 (68.3%) | 46/55 (83.6%) | 7/9 (77.7%) | 0.49 |
| D-Dimer ng/L | 2.6 (1.1-6.5) | 2.9(1.2-8.8) | 3.5(1.3-9.5) | 0.8 (0.5-1.2) | <0.001 |
| D- Dimer> 560ng/ml (0.56 ng/L) | 66/74 (89.2%) | 55 (91.7%) | 55/59 (93%) | 11/15 (73.3%) | 0.05 |
| C-reactive protein (mg/L) | 9.8 (4.5-18.9) | 13.5 (6.1-19.9) | 13.2 (5.9-19.1) | 7.3 (0.7-17.7) | 0.36 |
| C-reactive protein > 5mg/L | 47/63 (74.6%) | 41/50 (82%) | 42/54 (77.8%) | 5/9 (55.5%) | 0.16 |
| Procalcitonin ng/ml | 1.85 (0.5-6.3) | 1.8(0.47-3.84) | 2.1 (1.16-6.4) | 0.37 (0.12-6.17) | 0.12 |
| Procalcitonin > 0.15ng/ml | 55/62 (88.7%) | 43/50 (86%) | 47/51 (92%) | 8/11 (72.7%) | 0.1 |
| Albumin< 3g/dl | 45/71 (63.3%) | 36/56 (64.3%) | 40/57 (70.2%) | 5/14 (35.7%) | 0.02 |

<sup>1</sup>N in the column heading was used to calculate proportions if not otherwise specified. For variables with missing data, denominators are reported in each cell.
<sup>2</sup>Excluded: cancer, immunodeficiency, neuromuscular, chronic respiratory disease except asthma, congenital cardiopathies, chronic kidney failure.<sup>3</sup>GIS: Gastrointestinal symptoms, includes abdominal pain, diarrhea, emesis.<sup>4</sup>Trisomy 21 (n=1), Post recent appendectomy (n=2), hypothyroidism (n=2), undernutrition (n=1).<sup>5</sup>Lymphopenia: (<2 years: <4000, 2-3 years: < 3000, >4 years: <1500 cells/mcl)
\*O one-sided Fisher exact test/ Mann-Whitney test for comparison between cluster 1 and cluster 2.

In addition to fever, the most common clinical manifestations were cough (52%), conjunctivitis (50.6%), abdominal pain (51%), rash (48%), headache (45.3%), and diarrhea (44%). The median (IQR) duration of symptoms before admission was 7 (4–9) days. About 51% had lower respiratory symptoms, 85% had pulmonary infiltrates on radiograph or computed tomography, and 65.3% required oxygen administration with a non-rebreathable mask, high flow nasal cannula, or mechanical ventilation. Vasopressor support was used in 52% of the cases, and about 40% had a left ventricular ejection fraction <55% on echocardiography. Median values for markers of inflammation, coagulation, and system damage are summarized in Table 1.

Three subgroups were analyzed independently. In the first subgroup (n = 60), 15 cases with severe underlying disease were excluded. Clinical manifestations and outcomes did not differ significantly from those observed in the total sample (Tables 1 and 2).

**Table 2.**
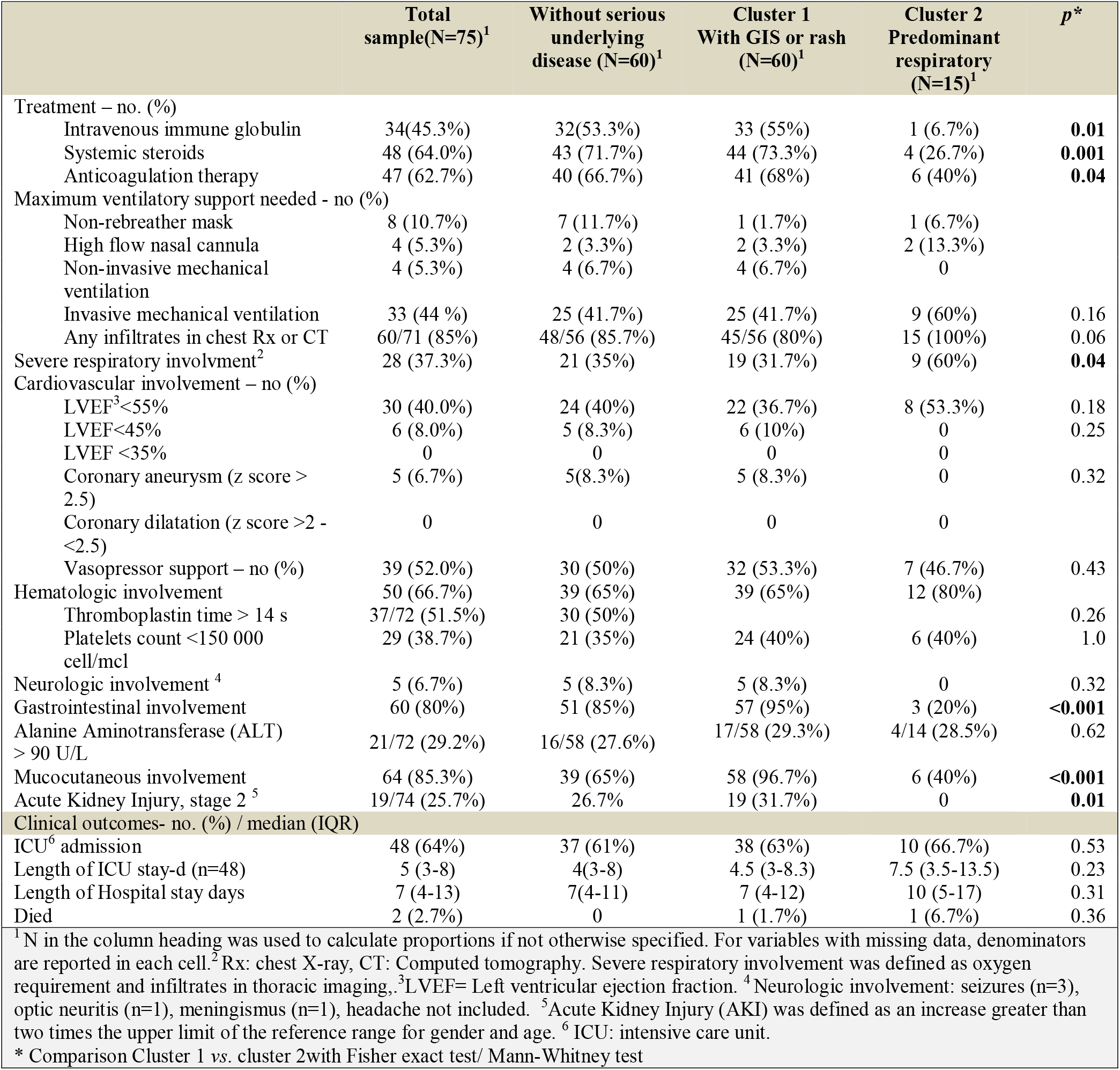
Treatment and outcomes of cases with PIMS.

The second (n = 60) and third (n = 15) subgroups were derived from LCA which identified two classes of patients: cluster 1 (n = 60) comprised of patients with rash and/or gastrointestinal symptoms, and cluster 2 (n = 15) comprised of those with predominantly cardiorespiratory involvement, without a rash or gastrointestinal symptoms. Despite the small number of cases in cluster 2, some differences were evident: cluster 2 cases were more prone to having a positive RT-PCR test (100% vs 76%, p = 0.04) and less likely to have a positive serology test (0% vs 54%, p < 0.001). An underlying medical condition was also more frequent in cluster 2 (80% vs 36.7%, p = 0.003), especially a severe one (47% vs 13%, p = 0.008) such as cancer, neuromuscular disability, or a chronic respiratory condition (Table 1). Cluster 1 patients tended towards higher levels of inflammation markers, though the difference was statistically significant for only the neutrophil count and D-Dimer concentration.

With regards to treatment, 45.3%, 64% and 63% of the cases received intravenous immune globulin, systemic steroids, and anticoagulation therapy respectively, which was more frequent in the cluster 1 group (Table 2). For outcomes, two (2.7%) patients died and they both had severe underlying diseases; the first patient had a medulloblastoma and died two weeks after the antineoplastic chemotherapy dose while the second one had chronic kidney failure (20) and was on renal replacement therapy. One death occurred in each cluster group. About 44% of the cases needed mechanical ventilation, 52% required vasopressor support while 40% had a depressed ventricular function. (Table 2)

All cases had an echocardiographic evaluation during hospitalization. Five cases (6.7%) had coronary aneurysms and notably, all these cases were in cluster 1. No patient had a coronary diameter with a z-score between 2.0 and 2.5. Myocardial involvement (left ventricular ejection fraction < 55%) was observed in 30 (40%) of the cases. ICU admission occurred in 64% of the cases with a median (IQR) length of ICU stay of 5 (3–8) days and a length of hospital stay of 7 (4–13). There was a tendency of cluster 2 patients to remain longer in ICU and the hospital. (Table 2).

**The third epidemic wave started at the beginning of June 2021** (18) **and was attributed mainly to the Delta variant of SARS-CoV-2, this variant being identified in about 80% of sequenced samples at the peak of this wave in mid-July** (19). **Eleven of the analyzed PIMS cases occurred after June 1**^**st**^, **2021. An exploratory analysis to compare outcomes between cases presenting before and after June 1**^**st**^ **showed that eleven PIMS cases occurred in the later period (Figure 1). A lower frequency of UTI admission was observed in this later period (36.4 vs 68.8%, p =0.05), and no deaths**.

## 4 Discussion

To our knowledge, the present study described the largest cohort of patients with PIMS in the Mexican population to date. Most clinical characteristics and outcomes were similar to those described in large international reports. The median age of our sample was higher than most of these previous reports. The prevalence of underlying medical conditions, including obesity, was similar to the reported data in the United States and Latin America (10,16,21–23), except for cancer which was relatively more frequent in our total sample, reflecting the characteristics of the population regularly attending the study site. Cardiac dysfunction, vasopressor requirements, rash, gastrointestinal symptoms, ICU admissions, and lengths of hospital and ICU stay were comparable to those reported elsewhere. The death rate of 2.7% (binomial 95% confidence interval: 0.3%–9.3%) was also consistent with previous reports (3,5,10,14,21,24).

Our population had a relatively high frequency of lower and serious respiratory involvement in comparison with most of the other studies. Besides, in our sample, serology was performed in only 55% of the cases, so the evidence of SARS-CoV-2 infection in our population was mostly by RT-PCR (80% positivity rate). These findings point toward a higher proportion of acute severe SARS-CoV-2 infection vs a theoretical late-onset post-infectious syndrome compared to other case series where respiratory involvement and RT-PCR positive rate was lower (3,5,10,14,21). Current PIMS diagnostic criteria do not exclude acute severe COVID-19; this overlap between both clinical presentations has been previously discussed (10,16).

On the other hand, the frequency of coronary aneurysms in our study was relatively low (6.7%). The frequency of aneurysms varied, ranging from 6.7% to 23.3% in previous studies (14,21,22,24). This probably reflects the lack of specificity in the PIMS case definition criteria and the variability of the cut-point for some inflammatory markers used in different reports. Most case series are derived from active or passive surveillance, either retrospectively or prospectively. Many of them do not inform a cut-point for inflammatory markers concentration, or clinical symptoms and the upper limit of the normal reference range may have been used. This probably explains the high frequency of coronary aneurysms (23.3%) found by Flood et al., in the United Kingdom. Their study had a very high cut-point of 100mg/L for C-reactive protein and required ‘acute abdomen’ and very specific dermatological findings, in addition to ‘abdominal pain’ or ‘rash’ as criteria for gastrointestinal or mucocutaneous involvement respectively (14). Stringency in diagnostic criteria automatically excludes a high proportion of potential cases, thus ensuring a more homogeneous and severely affected population.

**While clinical and epidemiological studies point toward the existence of at least two different clinical presentations of severe illness associated with SARS-CoV-2 infection (i.e**., **acute severe COVID-19 vs. PIMS)** (10,15,16,25,26), **a clear-cut distinction between the clinical and physio-pathological features of each one has remained elusive. On the one hand, severe COVID-19 is supposed to be the expression of an acute pulmonary infection with high viral loads; on the other hand, PIMS is conceived as a post-infectious syndrome with an exacerbated immune activation** (9,27,28). **Regarding physiopathology, some studies have identified different immunological profiles between pediatric acute COVID-19 and PIMS** (29,30)**; however, these studies fail to include groups of patients with severe-acute COVID-19 and of SARS-CoV-2 infected patients with severe coexisting comorbidities. Our study does include these two last groups of cases, which led to a more pronounced overlap and effacement between groups**.

Although ours was a small sample to make any inference, it supports the increasing awareness about the existence of different clinical phenotypes within positive PIMS criteria. Patients included in our cluster 1 were characterized by the presence of rash or gastrointestinal symptoms, with higher levels of some inflammatory markers. Custer 2 cases had a clinical presentation with predominant respiratory manifestations as described in severe acute COVID-19.

**Some researchers have tried to define subgroups within patients who fulfill PIMS criteria. We tested several LCA-derived models using factors described in previous attempts to classify cases. (14–16), and finally selected the current model which includes three factors (i.e**., **rash, gastrointestinal involvement, and severe respiratory involvement) as the one with the best balance between fit (i.e**., **Bayesian Information Criterion score) and parsimony**. Godfred-Cato et al. (15), and later Geva et al. (16), identified three classes of patients: 1) a seriously ill group with significant cardiovascular involvement, multiple organ dysfunction, a pronounced elevation of inflammatory markers with a tendency of positive serology for SARS-CoV-2, 2) a group with predominantly respiratory involvement and high RT-PCR positivity rate that resembles our cluster 2 group and 3) a group with predominantly Kawasaki-like mucocutaneous findings. Meanwhile, Flood et al. also identified three PIMS phenotypes using cluster analysis: 1) PIMS with Kawasaki disease-like presentation, 2) PIMS with findings similar to Kawasaki disease and shock and 3) PIMS without any of these features (14). Accordingly, consensus-derived clinical recommendations have been generated (31). **Of note, Flood specified very high cutting points of laboratory and clinical signs to qualify as a PIMS, thus getting a more homogeneous sample and with higher levels of inflammation than we did. This circumstance and a bigger sample size let them classify cases in more categories and with subtler differences than we did**.

In all three cluster analyses, coronary artery aneurysms were more frequent in groups with Kawasaki-like features and higher levels of inflammatory markers, which corresponds to cluster 1 in our cluster analysis. Notably, none of the five aneurysm cases in our sample had serious underlying diseases and all of them belonged to cluster 1 (Mucocutaneous/gastrointestinal predominant symptoms).

The role of inflammation in the spectrum of severe illness associated with SARS-CoV-2 infection should be deeply studied, including its role in myocardial dysfunction since it is a frequent complication both in adults with COVID-19 and children with PIMS (32). **The resemblance of PIMS with Kawasaki disease and the notion of a dysregulated immune activation has led to the utilization of a similar therapeutic approach with intravenous immune globulin and glucocorticoids** (33), **as well as the presumption of the possible effectiveness of immune modulators such as tumor necrosis factor, interleukin-1, and interleukin-6 inhibitors**.

A limitation of this study, as in others, is the method for identification of included cases for analysis, which depended on spontaneous reports by treating clinicians. In the future, a prospective multicentric cohort study with clear-cut criteria for subclassification of every patient fulfilling the PIMS broad clinical criteria after SARS-CoV-2 infection would provide more information on the outcomes and possible associated factors.

**Other limitations of the study are those inherent to the retrospective design which bias results towards the identification of more severe cases and to the lack of some data in the clinical files. It would have been desirable to have had the SARS-CoV-2 serology for every case as well as myocardial damage biomarkers. On the other hand, one of the strengths of the study is the availability of an echocardiographic evaluation of almost all patients, since heart involvement is one of the most feared complications associated with SARS-CoV-2 infection in children**.

## 5 Conclusions

The epidemiology of PIMS cases observed in our center is comparable to those reported elsewhere, although our study included a high proportion of patients with an acute respiratory phenotype. Coronary aneurysms were uncommon and usually present in previously healthy patients with rash or gastrointestinal symptoms.

It might be important to differentiate the clinical subtypes diagnosed as PIMS under the umbrella of CDC and WHO case definition criteria since probably not all of them benefit from the same therapeutic interventions.

The death rate paralleled previous reports and occurred in patients with severe underlying diseases.

This study agrees with other researchers that in the context of case definition criteria for COVID-19, at least two groups of patients can be identified according to their clinical presentation. Standardized criteria for subclassification of PIMS(28) cases need to be developed, since different phenotypes might have different pathophysiology, different outcomes, and possibly require different management. Clinical trials are needed to evaluate therapeutic interventions for different PIMS phenotypes.

## Data Availability

All data produced in the present study are available upon reasonable request to the authors

## 7 Author Contributions

All authors contributed to the study plan and design. RGG, MLMY, RNJJ, VBOL, CDB collected the data. HMG coordinated the research team. MFCP and NGG performed the data analysis and wrote the first draft of the manuscript. All authors reviewed and approved the final manuscript.

## 9 Data Availability Statement

The datasets generated for this study can be requested to corresponding authors.

---

**Chart 1.**
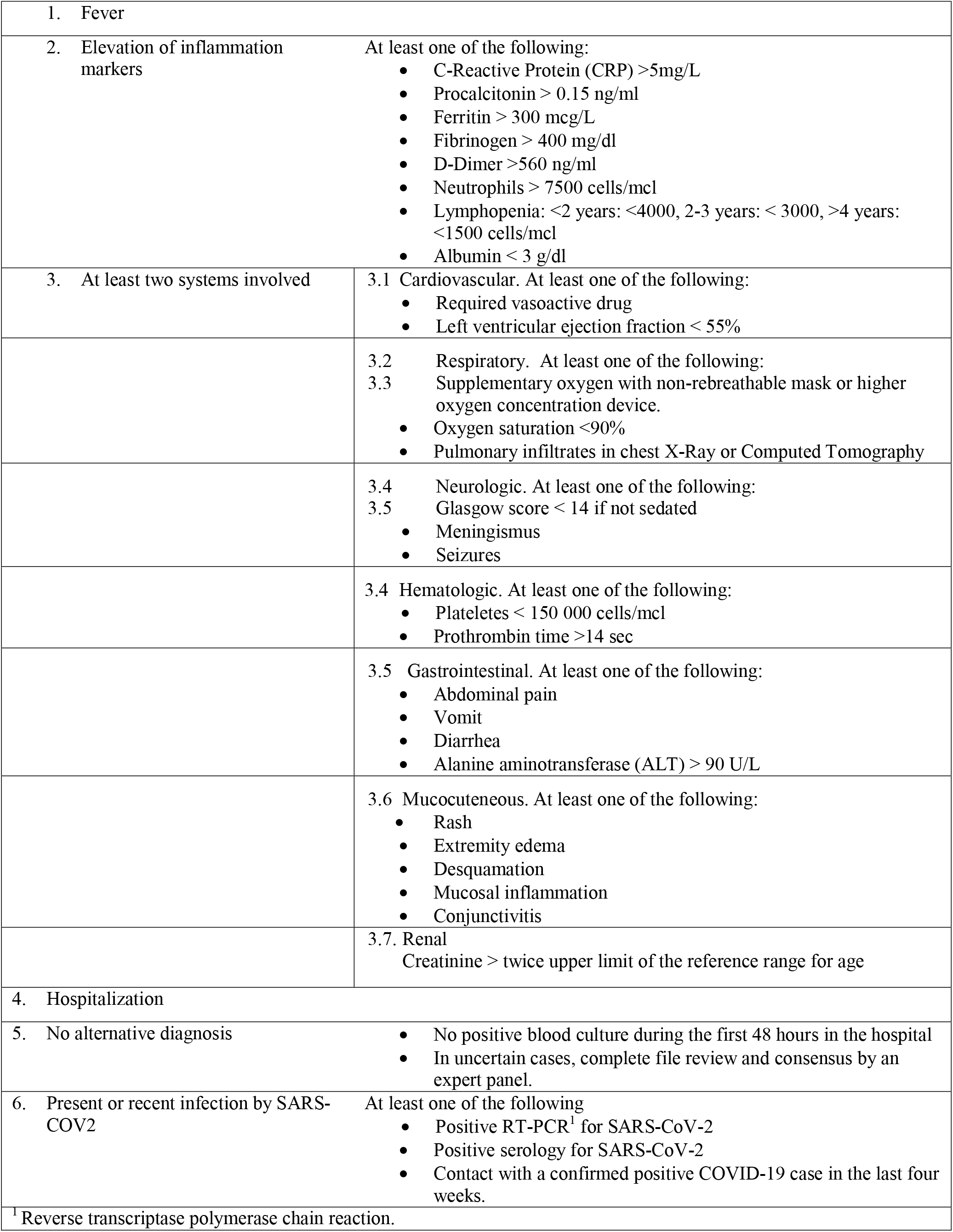
Case definition criteria for PIMS.

| Chart 1. Case definition criteria for PIMS |  |
| --- | --- |
| 1. Fever |  |
| 2. Elevation of inflammation markers | At least one of the following:<br>• C-Reactive Protein (CRP) >5mg/L<br>• Procalcitonin > 0.15 ng/ml<br>• Ferritin > 300 mcg/L<br>• Fibrinogen > 400 mg/dl<br>• D-Dimer >560 ng/ml<br>• Neutrophils > 7500 cells/mcl<br>• Lymphopenia: <2 years: <4000, 2-3 years: < 3000, >4 years: <1500 cells/mcl<br>• Albumin < 3 g/dl |
| 3. At least two systems involved | 3.1 Cardiovascular. At least one of the following:<br>• Required vasoactive drug<br>• Left ventricular ejection fraction < 55% |
|  | 3.2 Respiratory. At least one of the following:<br>3.3 Supplementary oxygen with non-rebreathable mask or higher oxygen concentration device.<br>• Oxygen saturation <90%<br>• Pulmonary infiltrates in chest X-Ray or Computed Tomography |
|  | 3.4 Neurologic. At least one of the following:<br>3.5 Glasgow score < 14 if not sedated<br>• Meningismus<br>• Seizures |
|  | 3.4 Hematologic. At least one of the following:<br>• Plateletes < 150 000 cells/mcl<br>• Prothrombin time >14 sec |
|  | 3.5 Gastrointestinal. At least one of the following:<br>• Abdominal pain<br>• Vomit<br>• Diarrhea<br>• Alanine aminotransferase (ALT) > 90 U/L |
|  | 3.6 Mucocutaneous. At least one of the following:<br>• Rash<br>• Extremity edema<br>• Desquamation<br>• Mucosal inflammation<br>• Conjunctivitis |
|  | 3.7. Renal<br>Creatinine > twice upper limit of the reference range for age |
| 4. Hospitalization |  |
| 5. No alternative diagnosis | • No positive blood culture during the first 48 hours in the hospital<br>• In uncertain cases, complete file review and consensus by an expert panel. |
| 6. Present or recent infection by SARS-COV2 | At least one of the following<br>• Positive RT-PCR<sup>1</sup> for SARS-CoV-2<br>• Positive serology for SARS-CoV-2<br>• Contact with a confirmed positive COVID-19 case in the last four weeks. |
| <sup>1</sup> Reverse transcriptase polymerase chain reaction. |  |

## References

1. Riphagen S, Gomez X, Gonzalez-Martinez C, Wilkinson N, Theocharis P. Hyperinflammatory shock in children during COVID-19 pandemic. Lancet. 2020;395(10237):1607–8.

2. Cheung EW, Zachariah P, Gorelik M, Boneparth A, Kernie SG, Orange JS, et al. Multisystem Inflammatory Syndrome Related to COVID-19 in Previously Healthy Children and Adolescents in New York City. JAMA - J Am Med Assoc. 2020;324(3):294–6.

3. Whittaker E, Bamford A, Kenny J, Kaforou M, Jones CE, Shah P, et al. Clinical Characteristics of 58 Children with a Pediatric Inflammatory Multisystem Syndrome Temporally Associated with SARS-CoV-2. JAMA - J Am Med Assoc. 2020;324(3):259–69.

4. Licciardi F, Pruccoli G, Denina M, Parodi E, Taglietto M. SARS-CoV-2 – Induced Kawasaki-Like Hyperin fl ammatory Syndrome_J: A Novel COVID Phenotype in Children. Pediatrics. 2021;146(2):1–5.

5. Dufort EM, Koumans EH, Chow EJ, Rosenthal EM, Muse A, Rowlands J, et al. Multisystem Inflammatory Syndrome in Children in New York State. N Engl J Med. 2020;383(4):347–58.

6. Assessment RR. European Centre for Disease Prevention and Control Country Experts. Rapid Risk Assessment: Paediatric inflammatory multisystem syndrome and SARS-CoV-2 infection in children. https://www.ecdc.europa.eu/sites/default/files/documents/covid-19-risk-assessment. 2020;(May):1–18.

7. HAN Archive - 00432 | Health Alert Network (HAN) [Internet]. [cited 2021 Dec 14]. Available from: https://emergency.cdc.gov/han/2020/han00432.asp

8. Royal College of Paediatrics and Child Health. Guidance paediatric multisystem inflammatory syndrome temporallly associated with Cov-19. R Coll Paediatr Child Heal. 2020;1–6.

9. Jiang L, Tang K, Levin M, Irfan O, Morris SK, Wilson K, et al. COVID-19 and multisystem inflammatory syndrome in children and adolescents. Lancet Infect Dis. 2020;20(11):e276–88.

10. Feldstein LR, Tenforde MW, Friedman KG, Newhams M, Rose EB, Dapul H, et al. Characteristics and Outcomes of US Children and Adolescents with Multisystem Inflammatory Syndrome in Children (MIS-C) Compared with Severe Acute COVID-19. JAMA - J Am Med Assoc. 2021;325(11):1074–87.

11. Son MBF, Murray N, Friedman K, Young CC, Newhams MM, Feldstein LR, et al. Multisystem Inflammatory Syndrome in Children — Initial Therapy and Outcomes. N Engl J Med. 2021;385(1):23–34.

12. Davies P, Evans C, Kanthimathinathan HK, Lillie J, Brierley J, Waters G, et al. Intensive care admissions of children with paediatric inflammatory multisystem syndrome temporally associated with SARS-CoV-2 (PIMS-TS) in the UK: a multicentre observational study. Lancet Child Adolesc Heal. 2020;4(9):669–77.

13. Payne AB, Gilani Z, Godfred-Cato S, Belay ED, Feldstein LR, Patel MM, et al. Incidence of Multisystem Inflammatory Syndrome in Children Among US Persons Infected With SARS-CoV-2. JAMA Netw Open [Internet]. 2021 Jun 1 [cited 2021 Dec 14];4(6):e2116420–e2116420. Available from: https://jamanetwork.com/journals/jamanetworkopen/fullarticle/2780861

14. Flood J, Shingleton J, Bennett E, Walker B, Amin-Chowdhury Z, Oligbu G, et al. Paediatric multisystem inflammatory syndrome temporally associated with SARS-CoV-2 (PIMS-TS): Prospective, national surveillance, United Kingdom and Ireland, 2020. Lancet Reg Heal - Eur [Internet]. 2021 Apr 1 [cited 2021 Dec 14];3. Available from: http://www.thelancet.com/article/S2666776221000521/fulltext

15. Godfred-Cato S, Bryant B, Leung J, Oster ME, Conklin L, Abrams J, et al. COVID-19-Associated Multisystem Inflammatory Syndrome in Children - United States, March-July 2020. MMWR Morb Mortal Wkly Rep [Internet]. 2020 Aug 14 [cited 2021 Dec 14];69(32):1074–80. Available from: https://pubmed.ncbi.nlm.nih.gov/32790663/

16. Geva A, Patel MM, Newhams MM, Young CC, Son MBF, Kong M, et al. Data-driven clustering identifies features distinguishing multisystem inflammatory syndrome from acute COVID-19 in children and adolescents. EClinicalMedicine [Internet]. 2021;40:09–10. Available from: https://doi.org/10.1016/j.eclinm.2021.101112

17. Linzer DA, Lewis JB. poLCA: An R package for polytomous variable latent class analysis. J Stat Softw. 2011;42(10):1–29.

18. COVID-19 [Internet]. [cited 2022 Feb 27]. Available from: https://covid19.healthdata.org/mexico/mexico-city?view=vaccinations&tab=trend

19. COVID-19 Data Explorer - Our World in Data [Internet]. [cited 2022 Feb 26]. Available from: https://ourworldindata.org/explorers/coronavirus-data-explorer

20. Ramaswamy A, Brodsky NN, Sumida TS, Comi M, Asashima H, Hoehn KB, et al. Immune dysregulation and autoreactivity correlate with disease severity in SARS-CoV-2-associated multisystem inflammatory syndrome in children. Immunity. 2021;54(5):1083–1095.e7.

21. Belay ED, Abrams J, Oster ME, Giovanni J, Pierce T, Meng L, et al. Trends in Geographic and Temporal Distribution of US Children with Multisystem Inflammatory Syndrome during the COVID-19 Pandemic. JAMA Pediatr. 2021;175(8):837–45.

22. States U, July M, Godfred-cato S, Bryant B, Leung J, Oster ME, et al. COVID-19–Related Multisystem Inflammatory Syndrome in Children. AAP Gd Rounds. 2020;44(3):30–30.

23. Torres JP, Izquierdo G, Acuña M, Pavez D, Reyes F. Multisystem inflammatory syndrome in children (MIS-C): Report of the clinical and epidemiological characteristics of cases in Santiago de Chile during the SARS-CoV-2 pandemic Juan. Int J Infect Dis. 2020;2020(100):75–81.

24. Antúnez-Montes OY, Escamilla MI, Figueroa-Uribe AF, Arteaga-Menchaca E, Lavariega-Saráchaga M, Salcedo-Lozada P, et al. COVID-19 and Multisystem Inflammatory Syndrome in Latin American Children: A Multinational Study. Pediatr Infect Dis J. 2020;40(1):1–6.

25. Flood J, Shingleton J, Bennett E, Walker B, Amin-Chowdhury Z, Oligbu G, et al. Paediatric multisystem inflammatory syndrome temporally associated with SARS-CoV-2 (PIMS-TS): Prospective, national surveillance, United Kingdom and Ireland, 2020. Lancet Reg Heal - Eur [Internet]. 2021 Apr 1 [cited 2021 Dec 14];3:100075. Available from: http://www.thelancet.com/article/S2666776221000521/fulltext

26. Mohsin SS, Abbas Q, Chowdhary D, Khalid F, Sheikh AS, Khan ZGA, et al. Multisystem inflammatory syndrome (MIS-C) in Pakistani children: A description of the phenotypes and comparison with historical cohorts of children with Kawasaki disease and myocarditis. PLoS One. 2021;16(6 June):1–13.

27. Vella LA, Rowley AH. Current Insights Into the Pathophysiology of Multisystem Inflammatory Syndrome in Children. Curr Pediatr Rep [Internet]. 2021;9(4):83–92. Available from: https://doi.org/10.1007/s40124-021-00257-6

28. Gruber C, Patel R, Trachman R, Lepow L, Amanat F, Krammer F, et al. Mapping systemic inflammation and antibody responses in multisystem inflammatory syndrome in children (MIS-C). medRxiv Prepr Serv Heal Sci. 2020;(January).

29. Sacco K, Castagnoli R, Vakkilainen S, Liu C, Delmonte OM, Oguz C, et al. Immunopathological signatures in multisystem inflammatory syndrome in children and pediatric COVID-19. Nat Med. 2022;

30. Consiglio CR, Cotugno N, Sardh F, Pou C, Amodio D, Rodriguez L, et al. The immunology of multisystem inflammatory syndrome in children with COVID-19. medRxiv. 2020;(January).

31. Harwood R, Allin B, Jones CE, Whittaker E, Ramnarayan P, Ramanan A V., et al. A national consensus management pathway for paediatric inflammatory multisystem syndrome temporally associated with COVID-19 (PIMS-TS): results of a national Delphi process. Lancet Child Adolesc Heal. 2021;5(2):133–41.

32. Eiros R, Barreiro-Pérez M, Martín-García A, Almeida J, Villacorta E, Pérez-Pons A, et al. Pericardial and myocardial involvement after SARS-CoV-2 infection: a cross-sectional descriptive study in healthcare workers. Rev Española Cardiol (English Ed. 2021;

33. McArdle AJ, Vito O, Patel H, Seaby EG, Shah P, Wilson C, et al. Treatment of Multisystem Inflammatory Syndrome in Children. N Engl J Med. 2021;385(1):11–22.

